# Regional changes in cellular and molecular pathology in children with end-stage dilated cardiomyopathy: Correlation with regional and global cardiac performance

**DOI:** 10.1101/2023.12.11.23299838

**Authors:** Andrea Pisesky, Ching Kit Chen, Mei Sun, John Duaz, Elizabeth Stephenson, John Coles, Mark K. Friedberg

## Abstract

**Background:** Paediatric dilated cardiomyopathy (DCM) carries a poor prognosis. We previously identified regional heterogeneity and patterns of left ventricular (LV) dysfunction that correlated with outcomes. In this project, we aimed to describe associations of regional myocardial performance with fibrosis and molecular signalling.

**Methods:** We prospectively studied children undergoing heart transplantation for DCM. Pre-transplant clinical and echocardiographic features were correlated with regional histological and molecular findings from explanted hearts. Ten LV and one right ventricular (RV) regions were assessed for fibrosis, myocyte area, and protein expression related to hypertrophy and fibrosis signalling (p38, ERK, phospho-JNK, phospho-GSK3β, SMA, cadherin, ILK), contractile function (myosin heavy chain), and calcium handling (SERCA2a, phospho-CamKII, phospholamban [PLN], and phospho-PLN).

**Results:** Eight children were included [median age 2.0years (0.3–15.1years)], of whom six required mechanical circulatory support. There was no difference in fibrosis burden or myocyte area between LV segments, and between ventricles. LV dimensions, ejection fraction and diastolic parameters were not related to fibrosis, myocyte area or molecular signalling. Tricuspid annular systolic plane excursion was related to myocyte volume (r=0.89, p<0.01). There was an inverse relationship between fibrosis and segmental longitudinal strain for LV basal and mid-posterior segments (basal posterior, r=0.96, p<0.01; mid-posterior, r=0.74, p=0.05). Global longitudinal strain was related to expression of ILK (r=0.78, p=0.02) and SERCA2a (r=0.71, p=0.04).

**Conclusion:** In paediatric end-stage DCM, regional cardiac function is associated with interstitial fibrosis and expression of calcium-cycling and contractile proteins. Phenotypic and molecular expression is variable. The RV shows similar injury and protein expression to the LV despite better myocardial function. These findings suggest that even with severely reduced LV function, paediatric DCM is a highly heterogeneous disease involving both ventricles.

## INTRODUCTION

Dilated cardiomyopathy (DCM) carries poor prognosis in children with 1- and 5- year rates of death or transplantation of 35% and 49%, respectively^1^. DCM manifests as a dilated left ventricle (LV) with profoundly reduced function. Right ventricular (RV) dysfunction is associated with worse clinical outcomes; however, it remains poorly characterized in children^2,3^. Although commonly perceived as a disease of global LV systolic dysfunction, echocardiographic studies have demonstrated heterogeneous regional wall mechanics^4^. It has been postulated that regional wall abnormalities contribute to progressive LV decline with the observation in our previous work that the most inefficient contraction patterns are associated with poorer outcomes^4^.

Regional myocardial function is impacted by a multitude of mechanical and molecular mechanisms. Animal models of electromechanical dyssyncrhony have demonstrated molecular abnormalities that mirror regional mechanics. Central among these are proteins involved in myocardial contractile function, calcium handling, hypertrophy, fibrosis and apoptosis^5,6^. While electromechanical dyssynchrony from electrical conduction delay is found in <10% of children with DCM^4^, regional abnormalities and increased mechanical dispersion are important contributors to global ventricular dysfunction^7,8^. However, the relationships between regional myocardial performance and the molecular mechanisms driving regional dysfunction in paediatric DCM are incompletely understood. This knowledge gap may contribute to the limited development of effective mechanism-based therapies and persistently high mortality. We hypothesized that abnormalities in regional myocardial performance correlate with regional injury and molecular signalling in children with DCM. Consequently, the aim of this study was to analyze the relation of regional myocardial mechanics and function to the expression of histological and molecular markers of myocardial fibrosis, hypertrophy and contractile function in paediatric DCM undergoing cardiac transplantation due to end-stage heart disease.

## METHODS

### Patient population

This was a prospective study of paediatric patients with DCM who underwent cardiac transplantation. Inclusion criteria were a LV end-diastolic diameter (LVEDD) Z score >2.0, LV ejection fraction (LVEF) <40% by two-dimensional echocardiography^9^ and cardiac transplantation. Exclusion criteria were cardiac pacing, congenital heart disease, atrial dysrhythmia and frequent ectopy precluding evaluation of consecutive sinus beats by echocardiography. Clinical, laboratory and echocardiographic variables within 30 days of transplantation were collected. The study was approved by the Hospital for Sick Children Research Ethics Board. Informed consent was obtained from the parents or legal guardians.

### Echocardiography

Echocardiography was performed using a Vivid 7 echocardiographic system (General Electric, WI, USA) following our comprehensive clinical functional protocol, in accordance to published guidelines including optimization of images for speckle tracking myocardial strain analysis^10,11^ (Supplemental text). LVEF was calculated using the modified Simpson biplane technique^12^. Study images were stored in raw Digital Imaging and Communications in Medicine format. Echocardiograms were analyzed offline using EchoPAC software (General Electric, WI, USA) by an observer blinded to the histology data. Standard segmentation was used to define the following segments: basal, and mid anteroseptal wall; basal and mid anterior wall; basal and mid anterolateral wall; basal and mid inferolateral wall; and basal and mid inferior wall^13^. This same segmentation was used for histological and molecular analysis.

### Myocardial tissue collection

Immediately after cardiac explantation, tissue samples were obtained from each of the aforementioned segments and cut into three specimens: two specimens were immediately snap-frozen in liquid nitrogen and stored at −80^ο^ Celcius for protein analysis. The third specimen was fixed in 10% neutral-buffered formaldehyde, processed and embedded in paraffin.

### Cardiac morphometry, collagen volume fraction and western blot

Transverse sections of the hearts (5-mm thick) were fixed in 10% formalin for 24-hours, dehydrated, and embedded in paraffin. Then, 4-µm microtome sections were prepared (Leica Microsystems A/S, Herlev, Denmark) and stained with Picrosirius red to visualize interstitial collagen, as previously described^14^. The proportional comparison of areas occupied by Picrosirius red–positive collagen versus the entire visual field was quantified morphometrically using automated planimetry (Adobe Photoshop CS2; Adobe, San Jose, CA)^14^. Five-micrometer cross-sections of myocardial tissue were cut and stained with hematoxylin-and-eosin to determine myocyte morphology (NIH ImageJ 1.6). The outline of 100-200 single myocytes were measured with CaseViewer images from hematoxylin and eosin–stained sections in each section. Western blot was complete as per laboratory protocol (Supplemental text).

### Statistical Analysis

Data are presented as mean with standard deviations or median with interquartile ranges, as appropriate. Comparisons of the structural and functional parameters between the different segments in patients were performed using one-way analysis of variance (ANOVA) with Bonferroni *post hoc* testing. Regression analysis was used to relate the extent of collagen, myocyte area, and protein expression to parameters of systolic and diastolic function (1 region/patient, using patients as the units of analysis). A *p*-value <0.05 defined statistical significance. The average values of interstitial fibrosis, myocyte area and protein expression in all the LV segments were used to assess the association with global LV function.

## RESULTS

### Patient Population

Eight patients (37.5% male) were included with a median age at diagnosis and transplantation of 1.0 years (range 0.4-17.0) and 2.0 years (range 0.3-15.1), respectively (**Table 1**). The underlying diagnosis was idiopathic DCM in six (75%) and anthracycline-induced cardiomyopathy (AIC) in two (25%) patients. All patients had advanced heart failure with Ross / New York Heart Association functional class III-IV. The mean QRS duration was 73ms (range 58-148ms) and the median heart rate 122bpm (range 70-143bpm). Of the eight subjects, six (63%) were previously on mechanical circulatory support: four (67%) with Berlin Heart EXCOR^®^ and two (33%) with HeartWare^®^ left ventricular assist device. The median duration of mechanical support was 95 days (range 59-666 days).

**Table 1.**

| Patient characteristics |  |
| --- | --- |
| Median age at diagnosis (years) | 1.1 (0.3 – 15.1) |
| Median age at transplant | 2.0 (0.3 – 15.1) |
| Gender (male) | 3.0 (38%) |
| Weight (kg) | 19.2 (5.6 – 47) |
| BSA (m<sup>2</sup>) | 0.7 (0.32 – 1.38) |
| Heart rate (bpm) | 136.0 (116 – 161) |
| Systolic blood pressure (mmHg) | 92.0 (75 – 114) |
| Diastolic blood pressure (mmHg) | 62.0 (51 – 81) |
| Mean arterial pressure (mmHg) | 71.0 (59 – 88) |
| QRS duration (ms) | 83.0 (58 – 148) |
| Aetiology |  |
| Idiopathic | 6 (75%) |
| Anthracycline-related | 2 (25%) |
| Medications |  |
| Diuretics | 7 (88%) |
| Antiarrhythmics | 1 (13%) |
| ACE inhibitors | 5 (63%) |
| β-blockers | 3 (38%) |
| Inotropes | 2 (25%) |
| Mechanical circulatory support | 6 (75%) |
| Median duration of MCS (days) | 248 (59 – 666) |

### Echocardiographic analysis

#### Left ventricular global and regional systolic function

All subjects had severely reduced global LV function (mean LVEF 22% (±9.8)) and severely dilated LVs (mean LVEDD Z score +7.1 (±4.1). Global and segmental longitudinal strain was severely reduced in all subjects with no regional differences observed in individual segments (**Table 2**, **Figure 1**). Diastolic function assessment was confounded by a short IVRT in four (50%) patients and a fused mitral valve E and A waves in four (50%) patients. The pulmonary vein A-wave was measurable in two (25%) patients. All subjects had mild to moderate RV dysfunction (mean RV FAC 33% (±16%)) and normal RV size (mean RV Z score 1.8 (±1.8)).

**Figure 1.**
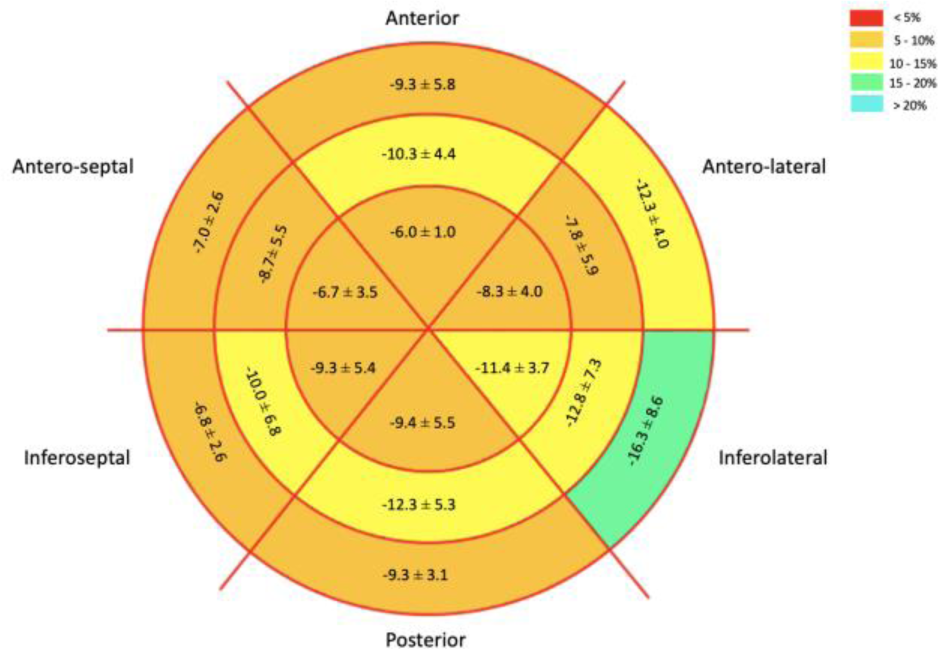
Bulls Eye of Left Ventricular Segmental Circumferential Strain Circumferential strain in mean with standard deviation as represented in a bulls eye.

**Table 2.**

| Parameter | N | Mean (SD) |
| --- | --- | --- |
| LV EF (%) Biplane | 8 | 21.9 (9.8) |
| LVEDd (cm) | 8 | 5.1 (0.6) |
| LVEDd z-score | 8 | 7.1 (4.1) |
| LVEDPW | 8 | 0.5 (0.2) |
| IVSD | 8 | 0.5 (0.2) |
| Global LV systolic LS (%) | 8 | -6.3 (2.7) |
| LV Doppler parameters |  |  |
| MV E (cm/s) | 8 | 109.7 (29.7) |
| MV A (cm/s) | 3 | 48.7 (17.2) |
| MV E/A | 3 | 2.1 (0.9) |
| DT (ms) | 3 | 98.0 (25.3) |
| IVRT (ms) | 3 | 54.4 (11.8) |
| PV S (cm/s) | 7 | 43.6 (18.8) |
| PV D (cm/s) | 7 | 46.7 (21.8) |
| LV lat e' (cm/s) | 8 | 7.4 (4.2) |
| LV lat a' (cm/s) | 6 | 2.8 (1.3) |
| LV lat s' (cm/s) | 8 | 3.6 (1.3) |
| IVS e' (cm/s) | 8 | 7.0 (2.1) |
| IVS a' (cm/s) | 6 | 4.3 (2.0) |
| IVS s' (cm/s) | 8 | 4.3 (1.6) |
| LV e/e' | 8 | 18.7 (9.3) |
| IVS e/e' | 8 | 16.5 (9.3) |
| RV parameters |  |  |
| RVEDd (cm) | 7 | 1.8 (0.8) |
| RVEDd z-score [mean (IQR)] | 7 | 1.8 (1.8) |
| RV FAC (%) | 8 | 32 (16) |
| TAPSE (mm) | 7 | 10.1 (4.7) |
| RV e' (cm/s) | 7 | 10.6 (6.3) |
| RV a' (cm/s) | 5 | 8.2 (6.0) |
| RV s' (cm/s) | 7 | 7.7 (3.8) |

#### Histological and Molecular Data

##### Interstitial fibrosis and myocyte area

All patients had histological and molecular analysis complete with a mean of five sections analysed per ventricular segment. There was no difference in global LV and RV fibrosis (LV 14.8±8.4%, RV 14.9±10.4%, p=0.56) or myocyte area (LV 1219±823µm², RV 1089±582µm², p=0.65). Children diagnosed at an older age had higher myocyte area (r=0.91, p=0.005, **Figure 2a**).

**Figure 2.**
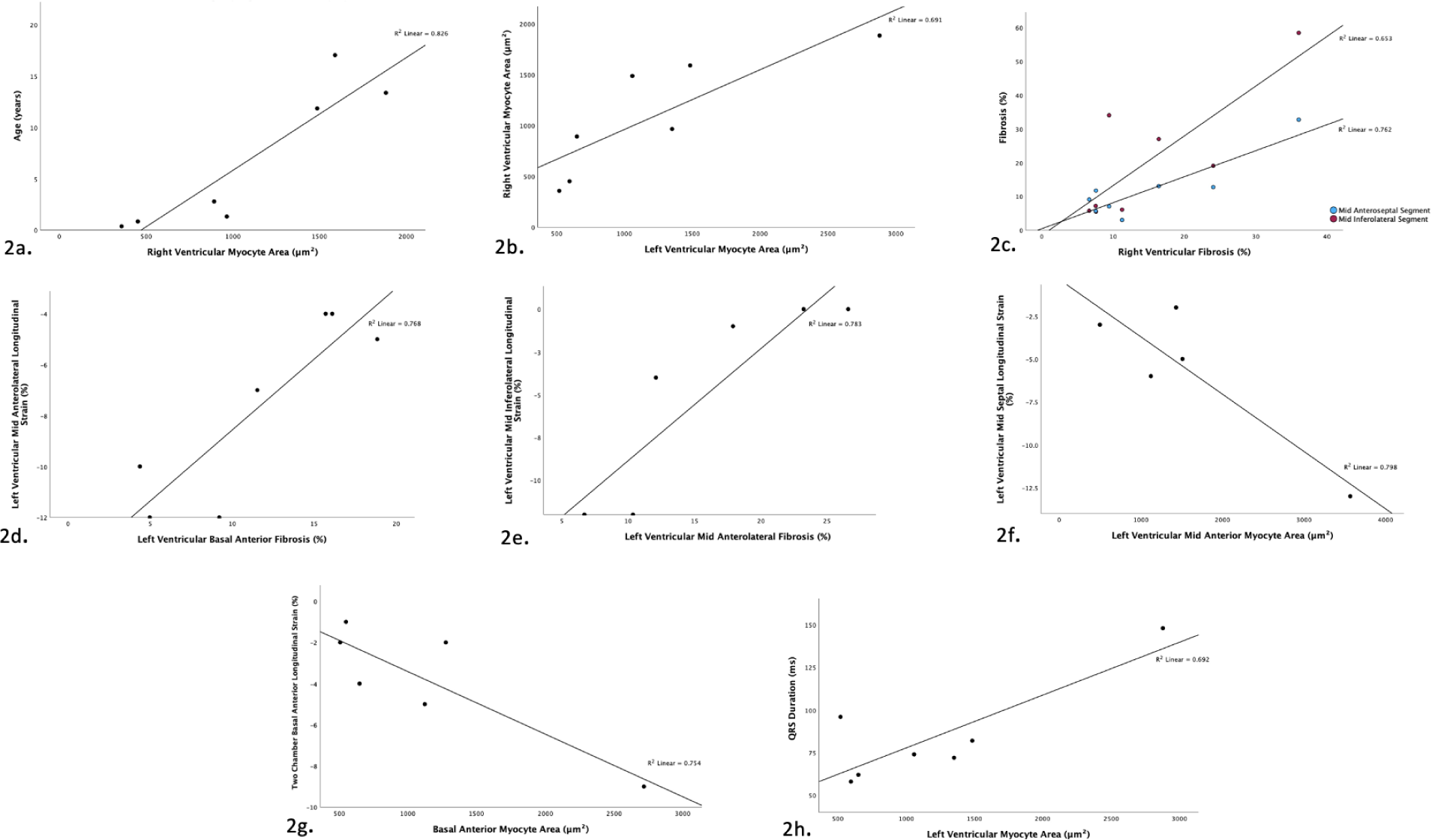
Correlations with myocyte area and ventricular fibrosis Scatter plot of age, in years, by right ventricular myocyte area; 2b - Scatter plot of right ventricular myocyte area by left ventricular myocyte area; 2c - Scatter plot of segmental fibrosis by right ventricular fibrosis; 2d - Scatter plot of basal ventricular fibrosis by left ventricular segmental longitudinal strain; 2e - Scatter plot of mid ventricular fibrosis by left ventricular segmental longitudinal strain; 2f - Scatter plot of left ventricular segmental myocyte area by segmental longitudinal strain; 2g - Scatter plot of segmental longitudinal strain by basal myocyte area; 2h - Scatter plot by QRS duration by left ventricular myocyte area.

There was a non-statistically significant trend of regional heterogeneity in fibrosis distribution within the LV myocardium (**Figure 3a**): the mid inferolateral segment had increased fibrosis and a large standard deviation as compared to the basal anteroseptal segment. There was markedly high variability in fibrosis burden between patients: patient one and three had significantly more fibrosis than patient four or five (**Figure 3b**). Segmental differences in interstitial fibrosis were present in three (37.5%) patients whereby one myocardial segment demonstrated more or less fibrosis as compared to the other segments (p<0.001 for patient one and four and p=0.013 for patient two). A similar trend was notable with myocyte size, whereby the basal inferior segment had more myocyte hypertrophy and higher variance (**Figure 3c**).

**Figure 3.**
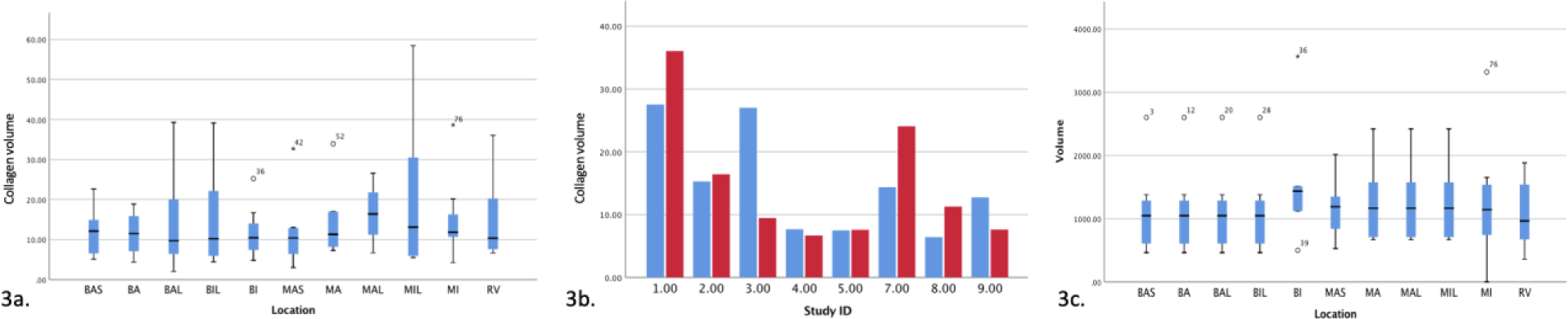
Interstitial fibrosis and myocyte area 3a - Collagen volume according to myocardial segment; 3b - Collagen volume according to patient, blue = left ventricular fibrosis, red = right ventricular fibrosis; 3c - Myocardial area according to myocardial segment BAS = basal anteroseptal, BA = basal anterior, BAL = basal anterolateral, BIL = basal inferolateral, MAS = mid anteroseptal, MA = mid anterior, MAL = mid anterolateral, MIL = mid inferolateral, RV = right ventricular.

There were no correlations between global LV or RV fibrosis and myocyte area (p=0.27). RV myocyte area correlated to LV myocyte area (r=0.83, p=0.01, **Figure 2b**). Additionally, RV fibrosis positively correlated to fibrosis in the LV mid anteroseptal and mid inferolateral segments (r=0.87, p=0.005 and r=0.81, p=0.015, respectively; **Figure 2c**).

There was an inverse and segmentally heterogenous, relationship between fibrosis and segmental longitudinal strain (e.g. in the basal (r=0.88, p=0.010) and mid inferolateral segments (r=0.89, p=0.0019), **Figure 2d and 2e**). Likewise, there was a correlation between myocyte area and segmental longitudinal strain in the mid anterior segment and basal septal segment (r=-0.90 p=0.041 and r=-0.84 p=0.038, **Figure 2f and 2g**), in that segments with myocyte hypertrophy had less segmental deformation.

Average myocyte area correlated with QRS duration (r=0.83, p=0.02, **Figure 2h**). RV myocyte area correlated with multiple parameters of global RV function including TAPSE (r=0.90, p=0.015, **Figure 4a**), TV S’ wave (r=0.92, p=0.003, **Figure 4b**) and A’ wave (r=0.90, p=0.005, **Figure 4c**.), and RV end diastolic dimension (r=0.95, p=0.001, **Figure 4d**). RV fibrosis did not correlate with any parameters of RV function, although there was a statistically non-significant trend to a negative correlation with TAPSE (RV FAC r=0.11, p=0.80; TAPSE r=-0.68, p=0.09; TDI TV S’ wave r=-0.57, p=0.14 and A’ wave r=-0.57, p=0.14; RV end diastolic dimension r=-0.07, p=0.87).

**Figure 4.**
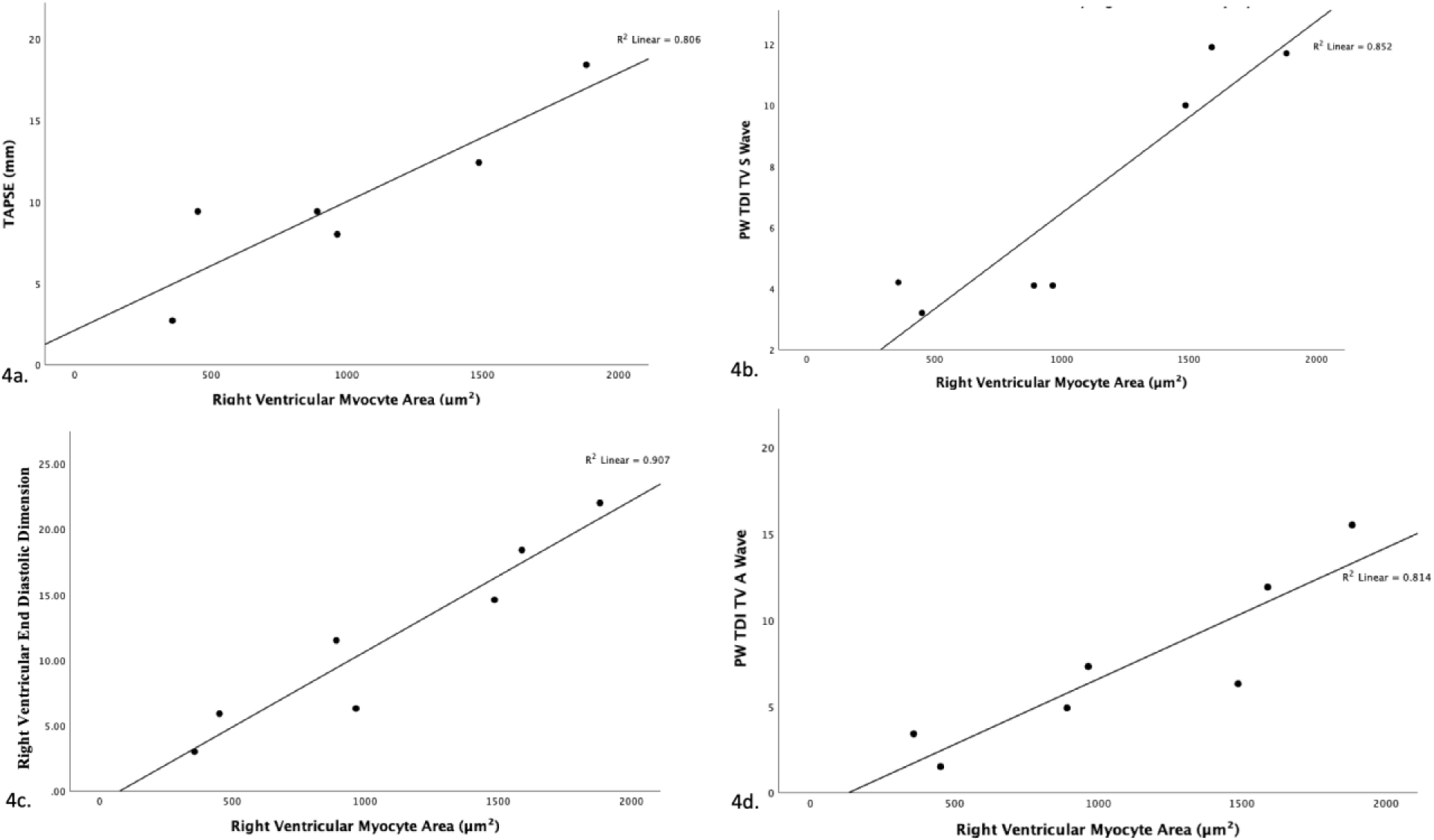
Correlations between right ventricular function and right ventricular myocyte area 4a - Scatter plot of TAPSE by right ventricular myocyte area; 4b - Scatter plot of PW TDI TV S wave by right ventricular myocyte area; 4c - Scatter plot of right ventricular end diastolic dimension by right ventricular myocyte area; 4d - Scatter plot of PW TDI TV A wave by right ventricular myocyte area

#### Protein expression

##### Expression of Proteins related to Contractility, Hypertrophy and Fibrosis

The expression of SMA, MYHC and αMHC was highest in the anterior and anteroseptal segments but overall variable between LV segments. Expression of cadherin and ILK was relatively homogenous between LV segments (**Figure 5a**). RV involvement was variable between proteins and patients. The expression of ILK, for example, was homogeneously expressed in both the LV and RV for all patients while the expression of MYH7 was higher in the RV in four patients and αMHC expression was variable between patients (**Figure 5b**). Correlations were observed between LV myocyte area and MYH7 expression (r=0.89, p=0.017, **Figure 6a**) and global αMHC expression (r=- 0.90, p=0.015, **Figure 6b**) such that higher expression of MYH7 *and* αMHC was associated with myocyte hypertrophy.

**Figure 5.**
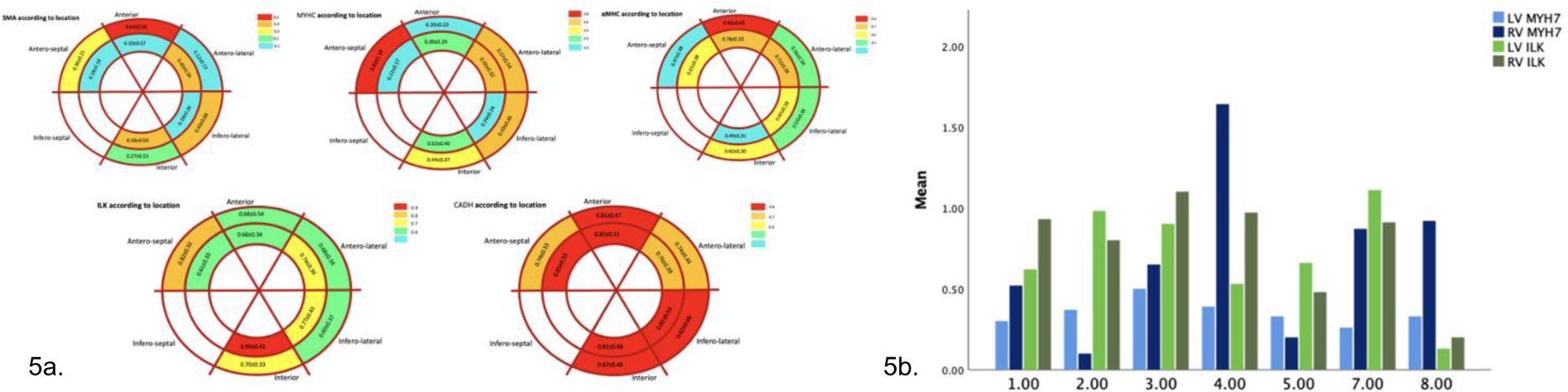
Expression of CADH, ILK, SMA, MYHC and αMHC 5a - Schematic representation of expression of CADH, ILK, SMA, MYHC and aMHC according to myocardial segment; 5b - Clustered bar mean of MYH7 and ILK protein expression by location and study ID

**Figure 6.**
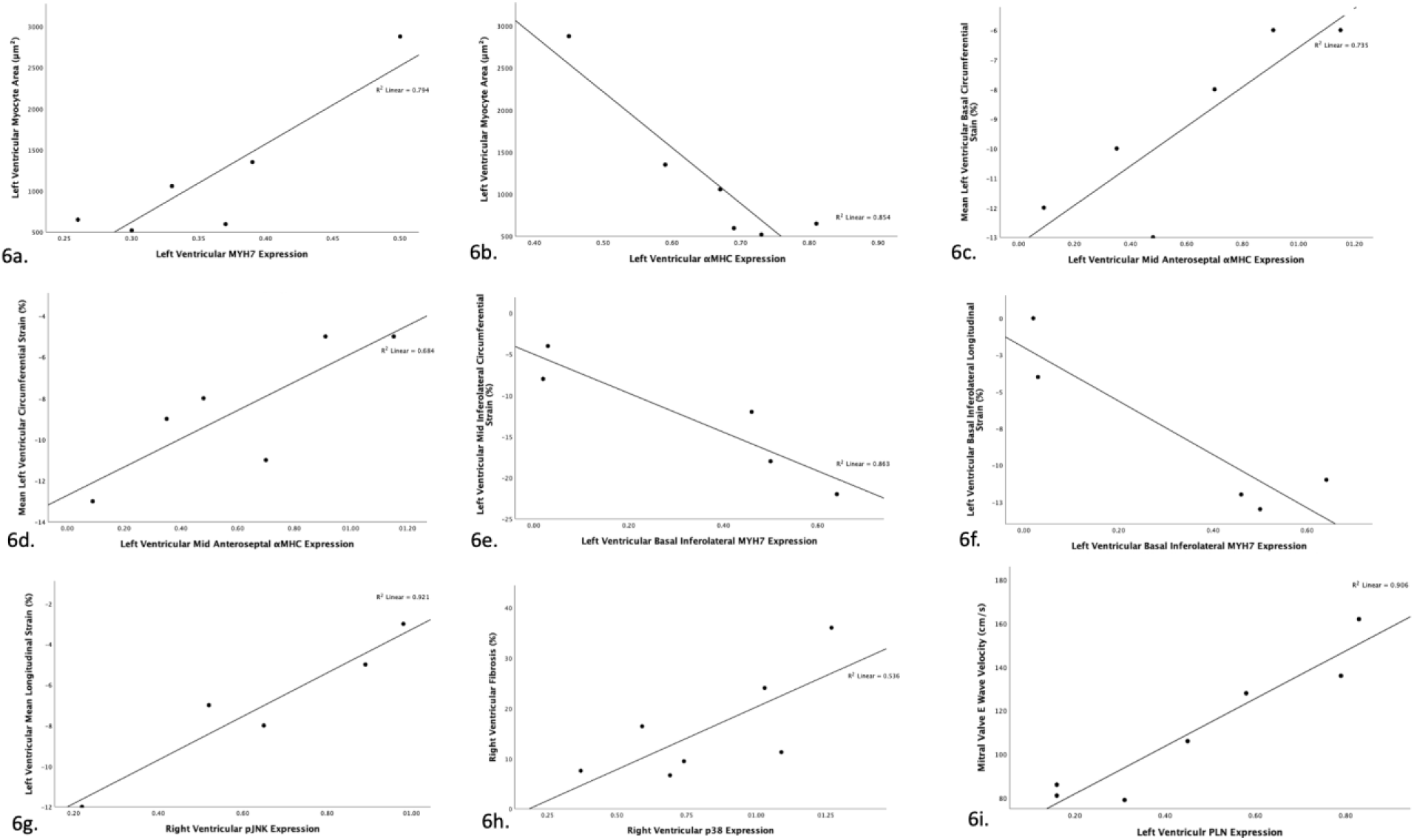
Molecular correlations 6a - Scatter plot of left ventricular myocyte area for left ventricular MYH7 protein expression; 6b - Scatter plot of left ventricular segmental strain by global left ventricular αMHC proteins expression; 6c - Scatter plot of left ventricular segmental strain by segmental αMHC protein expression; 6d - Scatter plot of circumferential strain by segmental αMHC protein expression; 6e - Scatter plot of segmental circumferential strain by segmental MYH7 protein expression; 6f - Scatter plot of Left ventricular segmental longitudinal strain by basal MYH7 protein expression; 6g - Scatter plot of left ventricular longitudinal strain by right ventricular pJNK protein expression; 6h - Scatter plot of right ventricular fibrosis by right ventricular p38 protein expression; 6i - Scatter plot of mitral valve E wave velocity by left ventricular PLN protein expression

##### Correlation of contractile proteins with segmental function

There were multiple strong correlations between segmental strain and segmental expression of contractile proteins: αMHC expression in the mid anterolateral segment correlated with basal and global circumferential strain (r=0.86, p=0.029 and r=0.83, p=0.04, **Figure 6c** and **Figure 6d**, respectively); MYH7 in the basal inferolateral septal area corresponded to circumferential strain in the mid inferolateral segment (r=-0.93, p=0.023, **Figure 6e**) and longitudinal strain in the basal inferolateral segment (r=-0.92, p=029, **Figure 6f**).

##### Expression of cell cycle activity, hypertrophy and fibrosis related proteins

The expression of p38 and ERK was homogenous throughout the LV and the RV. Conversely, there was heterogenous expression of p-JNK and p-GSK with increased pGSK expression in the basal inferior and mid inferolateral segments and increased p-JNK in the basal anterior and mid inferior segments (**Figure 7a**). The expression of p-GSK was particularly variable between patients, with subject 1 having much less expression than subject 2, for example, and subject 8 demonstrating a LV dominant pattern (**Figure 7b**). RV p-JNK expression inversely correlated with global longitudinal strain (r=0.96, p=0.01, **Figure 6g**.).

**Figure 7.**
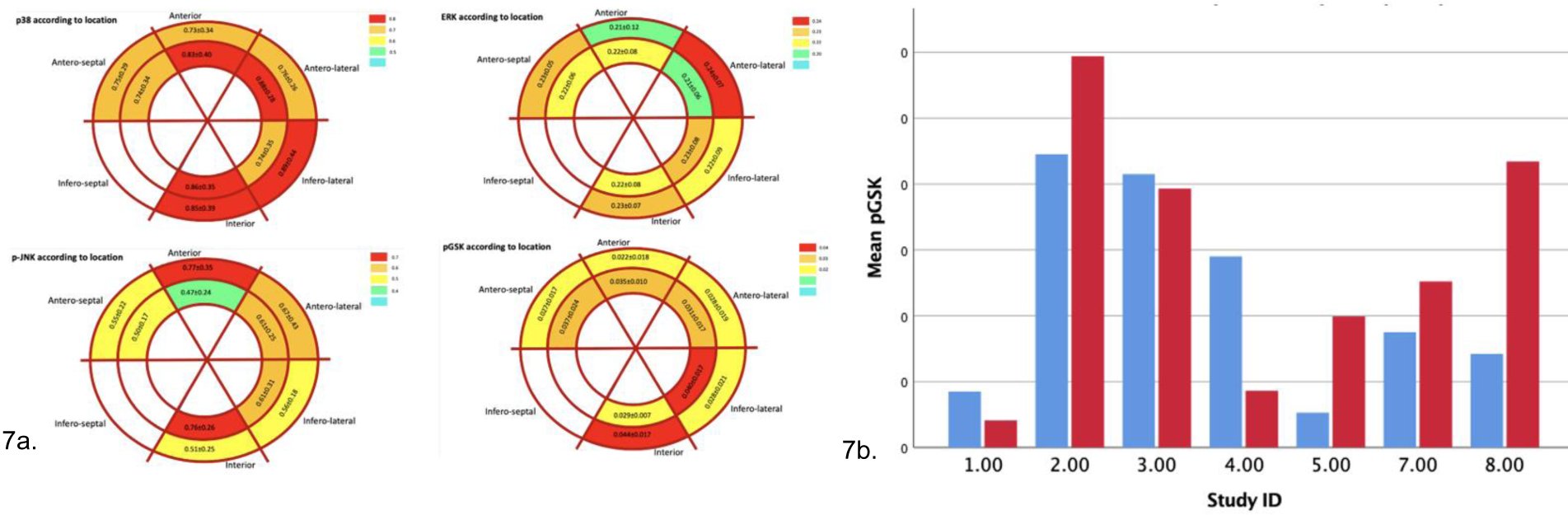
- Expression of p38, ERK, pJNK and pGSK 7a - Schematic representation of expression of p38, ERK, p-JNK and p-GSK; 7b - Expression of pGSK by study ID and location, blue = left ventricular, red = right ventricular.

##### Expression of calcium handling proteins

Calcium signalling proteins were heterogenously expressed between LV segments. For example, SERCA2a expression was relatively increased in the mid anterolateral and inferior segments and p-CamkII expression was relatively increased in the basal inferolateral segment (**Figure 8a**). There was a notable difference in calcium signalling between patients: subject 8 had less expression of all calcium signalling proteins while subject 7 had increased PLN expression as compared to peers (**Figure 8b**). There was no statistically significant difference in calcium protein expression between the RV and LV. As for other proteins, there was marked variability between patients. For example, patient 2 had an LV dominant PLN expression and patient 3 had a RV dominant Serca2a expression (**Figure 6h**). Calcium handing did not correlate to fibrosis, myocyte hypertrophy or markers of systolic function. Left ventricular expression of PLN correlated with the mitral valve E wave velocity (r=0.952, p<0.001, **Figure 6i**.).

**Figure 8.**
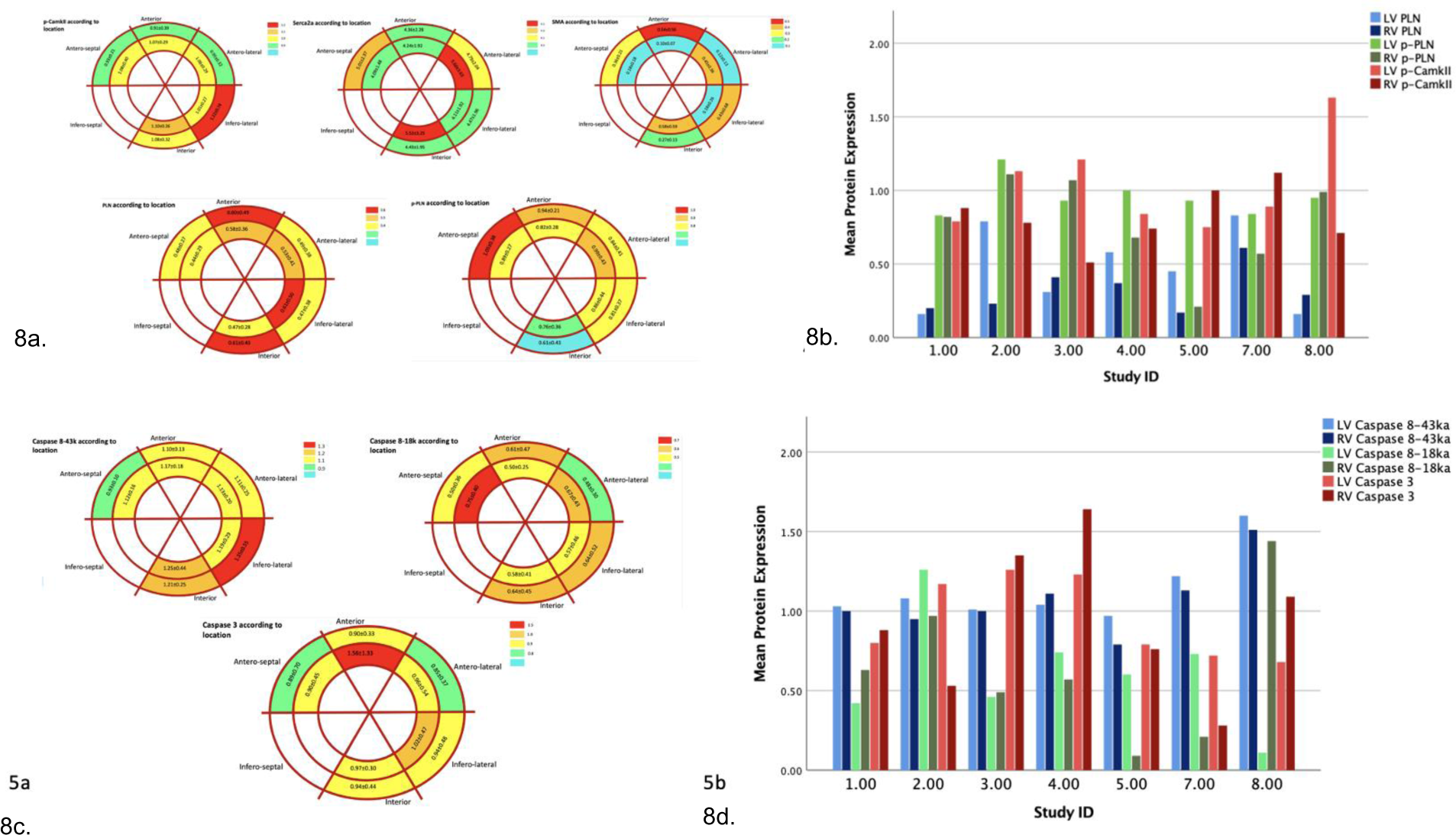
- Expression of p-CamkII, Serca2a, SMA, PLN, and p-PLN Left and right ventricular expression of PLN and p-PLN by study ID. 8c - Schematic representation of expression of Caspase-8 and Caspase-3; 8d - Left and right ventricular expression of Caspase-8 and Caspase-3 by study ID

##### Protein expression of apoptosis-related proteins

Apoptosis-related protein expression was heterogenous in the LV, with increased expression of caspase 8-43ka in the basal inferolateral segment, caspase 8-18ka in the mid anteroseptal segments and caspase 3 in the mid anterior segment (**Figure 8c**). Caspase 8-43ka and caspase 3 protein expression was similar in the LV and RV for all patients, whereas the expression of caspase 8-18ka protein in the LV and RV varied between patients: patient 8 had an RV dominant expression, patients 7 and 5 had LV dominant expression and the remainder biventricular expression (**Figure 8d**). There were no correlations between markers of apoptosis and myocyte hypertrophy or fibrosis, global systolic or diastolic function or ventricular volumes except RV fibrosis correlated with RV p38 expression (r=0.732, p=0.031, **Figure 6c**).

### Clinical correlations

Comparing patients with AIC and idiopathic DCM, there was no significant difference in LV fibrosis (AIC 10.1±3.7% vs DCM 16.4±9.1%, p=0.40) or myocyte area (AIC 1270±300µm² vs DCM 11198±996µm², p=0.93). The use of an ACE inhibitor medication was not associated with any difference in fibrosis (15.2±7.5% with ACE vs 14.2±11.5%, p=0.89) or myocyte area (1402±1065µm² with ACEi vs 976±421µm², p=0.55). Patients on MCS had higher caspase 8 expression levels in the entire LV (p=0.05), notably higher in the anterior basal regions. There were segmental differences for children on MCS whereby αMHC (p=0.04), MYH7 (p=0.001) and pJNK (p=0.01) protein expression was increase in the basal segments as compared to DCM patients not on MCS.

## DISCUSSION

DCM in children manifests with severe LV remodeling and severe LV systolic and diastolic dysfunction. RV function has received less attention and varies between patients. The regional functional and molecular findings in this cohort suggest that while abnormal expression of contractile proteins is related to regional deformation, other mechanisms including fibrosis signalling, cell cycle activity and calcium handling also correlate with abnormal cardiac mechanics. Although performed in a small number of patients, and although this cross-sectional study cannot establish causation, our results afford the following insights into end-stage paediatric DCM:

- Despite better function, the RV is significantly affected with molecular abnormalities similar in severity to those observed in the LV.
- Abnormal regional mechanics are associated with increased regional fibrosis.
- The relationships between molecular and functional abnormalities are heterogenous between patients and within LV segments of individual patients.
- Correlations between deformation and signalling pathways, particularly those related to fibrosis and hypertrophy, while heterogenous are higher at the ventricular base.
- ERK-MAPK signalling and hypertrophy-fibrosis proteins correlate with functional parameters more than calcium handling proteins.

### Fibrosis is prominent in the RV and LV

Exemplified by our cohort, fibrosis is an important pathologic response found in end-stage, non-ischaemic DCM^3,15^; an observation supported by single-cell RNA sequencing demonstrating activated fibroblasts in DCM patients^16^. The segmental pattern of fibrosis and hypertrophy was variable between our patients, perhaps reflecting different mechanical stresses and pathophysiologic response. While the normal LV is a geometrically symmetric and homogeneous chamber, our results confirm that DCM represents a wide spectrum of disease severity and histologic phenotypes. As in our prior studies, we believe the results of the current study supports the notion that dispersed or asymmetrical function and myocardial molecular remodeling between segments may stem from and/ or contribute to global ventricular inefficiency and hence dysfunction.

Previous paediatric studies have demonstrated that patients exhibit specific patterns of impaired deformation and that a heterogeneous pattern is predictive of adverse outcomes^4,7^. Given our cohort represented those patients with the poorest outcomes, we would have expected to primarily see a heterogenous deformation pattern; however, strain analysis demonstrated a homogenous pattern of impaired regional cardiac mechanics. It is possible that disease severity, the use of mechanical circulatory support or averaging results impeded the detection of statistically significant differences in peak regional strains. The results of this study, however, demonstrate the physiological consequences of abnormal regional mechanics in that there was an inverse relationship between the extent of interstitial fibrosis and segmental longitudinal strain: segments with increased fibrosis or myocyte area were observed to have lower deformation, a finding previously shown in adult DCM^17^. The observed relationship between histological abnormalities and segmental dysfunction despite the globally impaired function underscores the importance of understanding the mechanisms driving mechanotransduction in DCM^18^ as a basis for of mechanism-targeted therapy, such as Lysyl oxidase-like 2 targeted therapy in adult non-ischaemic heart failure^19^, dapagliflozin in diabetic cardiomyopathy^20^, and rapamycin in adriamycin-induced DCM^21^.

Although, RV involvement is well-recognized, DCM, is generally considered to be a ‘left-heart’ disease. RV fibrosis was highly prevalent in our cohort and similar in severity to that observed in the LV. This underscores that DCM is a biventricular disease. However, despite the universal presence of RV fibrosis, RV function was less impaired than LV function and there were no correlations between RV fibrosis and function, findings also reported in an adult DCM cohort^22^. This observation is valuable given that previous adult and paediatric DCM data supports the notion that RV dysfunction is predictive of outcomes such as transplantation and death^23^. The lack of significant RV dysfunction in our cohort may be explained by different threshold for transplantation or different mechanistic DCM phenotypes, such as the metabolic profibrotic phenotype whereby there is preserved RV function, universal midwall myocardial fibrosis and an association with diabetes and renal dysfunction^24^. Although tempting to link the underlying pro-fibrotic mechanism to a hyper-activated renin angiotensin state, adverse RV remodeling is a complex multifactorial maladaptive process involving structural, hemodynamic, histopathological, metabolic and genetic changes. Furthermore, mechanism-targeted therapy has the potential of preventing disease progression. For example, Angiotensin-1–7 ameliorates RV fibrosis and dysfunction in diabetic rats without correcting hyperglycemia^25^. The present study adds to our understanding of paediatric DCM phenotypes by identifying a subtype with diffuse RV fibrosis without profound dysfunction. These results should stimulate research into the pathological and prognostic significance of RV and LV fibrosis in paediatric DCM, especially as non-invasive assessment of fibrosis by magnetic resonance imaging is used in clinical prognostication of adult DCM^26^.

### The QRS duration is prolonged and associated with myocardial hypertrophy

QRS prolongation is recognized as an important characteristic of heart disease: present in end-stage heart failure^27^, predictive of mortality in ICD patients with heart failure^28^ and associated with regional wall motion abnormalities in myotonic dystrophy^29^. In parallel to observations from a rabbit model of heart failure^30^, QRS duration in our cohort correlated with myocyte hypertrophy; however, without any relationship to parameters of global systolic dysfunction or deformation. The resulting conduction velocities have been shown to be dynamic^30^, associated with disease severity^31^, and postulated to be implicated in the development of arrhythmias. We previously showed that while mechanical dispersion is common, frank electromechanical dyssynchrony is present in <10% of the paediatric DCM population. Thus, regional histological and molecular heterogeneity in our study does not likely originate from electromechanical dyssynchrony but may be the cause and/ or effect of increased mechanical dispersion.

### Relation between molecular and functional abnormalities

In dilated failing hearts, multiple molecular pathways are activated^32,33^ of which we sampled those associated with fibrosis, hypertrophy, calcium handling, apoptosis, and cell cycle activity. Overall, the heterogeneous molecular expression between patients and between LV segments, makes it challenging to suggest specific therapeutic targets. In most patients, the LV base appeared to have higher expression of proteins related to fibrosis and calcium signalling. This observation is consistent with higher wall stress at the base of the heart as observed in pressure-loaded RVs, associated with basal hypertrophy and apical sparing^34^. Interestingly, p-JNK and p-GSK expression was more upregulated at the ventricular base which may relate to lower strain and higher fibrosis in this area. These factors can be associated with pro-fibrotic TGFβ non-canonical signalling and are upregulated in RV fibrosis^35^. JNK signalling also plays a role in fibroblast proliferation and activation. Activation of JNK1, by TGFβ1 and to a lesser extent by Wnt signalling, results in activation of the AP-1 protein complex (consisting of c-jun and c-fos transcription factors) which promotes expression of cell-cycle regulator proteins such as cyclin D1^36^. JNK and p38 MAPK signalling can also mediate fibrosis through regulation of MMPs and TIMPs^37^; and matrix metalloproteinase inducer (EMMPRIN) expression is reportedly increased in DCM patients^38^. β-adrenergic stimulation of dominant-negative JNK rat ventricular cardiomyocytes showed reduced EMMPRIN expression^37^. Increased p38-MAPK activity has also been shown to mediate expression of c-fos transcription factor, and in turn, promote expression of MMPs in human cardiac fibroblasts^39^. Additionally, p38-MAPK can increase αSMA expression, a marker of fibroblast activation, via the SRF transcription factor^40^. Therefore, these pathways may, in part, explain αSMA expression and fibrosis in the LV anterior, inferior, and infero-lateral segments.

Increased p-GSK3β in our samples suggests activation of Wnt signalling in DCM. Serum Wnt5a levels correlate with worse RV dysfunction and mortality in DCM patients^41^. However, activation of Wnt signalling in adult DCM is often debated. One study showed decreased β-catenin levels in DCM patients, while others showed that in patients with idiopathic DCM and ischemic heart disease, increased myocardial β-catenin stabilization and nuclear translocation^42^. Concordant with the latter study, increased phosphorylation of GSK3β at the ser9 position is associated with β-catenin stabilization and nuclear translocation. Nuclear β-catenin can act as a transcriptional co-activator promoting the expression of fibrosis-related genes, including COL1A1 and POSTN, and genes that stimulate cardiac fibroblast proliferation^43,44^. In end-stage DCM patients Wnt5a expression may correlate with NFAT target gene activation^41^, via JNK activation^45^. This may suggest a potential connection between the two upregulated proteins found in our samples. Together, our results suggest a potentially important clinical role of Wnt signalling in LV and RV remodeling and fibrosis in paediatric DCM.

αMHC and MYH7 protein expression correlated with myocardial strain in particular segments: higher αMHC expression correlated with better deformation whereas higher MYH7 expression correlated with worse strain. These observations suggest that strain is associated with the action of contractile proteins, both being impacted by loading. In the healthy mammalian heart, αMHC, representing only 10% of the ventricular myosin heavy chain isoforms, is known to have a disproportionately large impact on contractile power production through fast ATPase activity^46,47^. In the pathophysiologic state of heart failure, functional alterations in αMHC expression, as seen in the genetic mutations associated with DCM as well as in our cohort, seem to importantly impact on cardiac contractility^47^.

We observed regional correlations between calcium handling proteins, such as phospho-PLN and SERCA2α, and abnormal mechanics. Although SERCA2α over-expression has been shown to restore contractility of cardiomyocytes in failing hearts^48,49^, increased expression in our cohort was associated with decreased longitudinal strain perhaps representing a maladaptive response. Although our ability to assess diastolic function was very limited, the correlations observed between calcium handling proteins and diastolic dysfunction are in line with mechanisms observed in hypertensive heart disease and hypertrophic cardiomyopathy^50^.

Our analysis demonstrated that all markers of apoptosis were heterogeneously expressed in the LV. Notably, there were no correlations between markers of apoptosis and myocyte hypertrophy or fibrosis with global systolic or diastolic function or ventricular volumes appreciating that these global parameters reflect the complex array of events in the failing heart.

## LIMITATIONS

Our cohort was biased to represent only those paediatric patients who underwent transplantation. A second limitation is the small sample size, precluding complex correlation assessment. Third, the RV was only sampled at one location and thus it was not possible to investigate regional right ventricular mechanics or protein expression. We did not have access to healthy control myocardium, which is difficult to obtain in children, and further difficult to match for age and sex. We intended using the RV sample as an internal control for LV findings. However, given the significant histological and molecular RV involvement, this was not appropriate.

## CONCLUSIONS

This study demonstrated association of regional cardiac function in children with DCM with interstitial fibrosis and expression of contractile and calcium-cycling proteins. Our results highlight that paediatric DCM is a biventricular disease characterized by prominent fibrosis and significant segmental myocardial heterogeneity. The relative preservation of RV function despite significant interstitial fibrosis and aberrations in protein expression may point towards a novel phenotype and warrants further investigation in relation to the functional and prognostic significance of fibrosis and myocyte hypertrophy in the RV versus the LV.

## Data Availability

All data referred to in this manuscript is available.

## Acknowledgments

We would like to thank Cameron Slorach, RDCS, who performed the echocardiography for this study.

